# Efficacy of Different Treatment Approaches (Insulin and Metformin) in Women with Gestational Diabetes Mellitus (GDM)

**DOI:** 10.1101/2024.01.09.24300942

**Authors:** Natalia Asatiani, Ramaz Kurashvili, Elena Shelestova

## Abstract

**Background and Purpose:** The pharmacotherapy options in patients with GDM, are insulin or oral antihyperglycemic agents. Insulin is the preferred medication for treating hyperglycemia, but in recent years, metformin has been increasingly used in the treatment of GDM.

**The aim:** to assess the efficacy of different treatment (insulin and metformin) in women with GDM.

**Methods:** Screening for GDM was performed in 2422 pregnant women and reveal GDM in 119 women (75-g OGTT was performed at 24-28 weeks of gestation). All patients started treated at 24 -29 weeks of gestations. The patients were divided into 2 groups (Gr.): Gr.1 - 68 patients, who treated with diet + insulin therapy Gr.2 - 51patients, who treated with diet + metformin.

**Results:** At the 2nd trimester HbA1c (%) levels vas for Gr.1 and 2: 6.7 (0.05), 6.4 (0.6) respectively; By term HbA1c levels statistically decreased in both groups, but we did not found statistical difference between groups. Women from Gr.2 gained less weight compared to women from Gr.1 (1.89±3.88 vs 4.53±3.67 kg; P=0.003). In Gr.1 percent of pre-eclampsia was (2,9%) and in Gr.2(3,9%) (P= 0,7773, OR – 1,33), we did not find statistical difference between groups. The incidence of preterm delivery before 37 weeks of gestation in Gr1 was lower, than in Gr.2 (P=0.7311, OR –1,33) and we also did not find a statistical difference between groups. Perinatal mortality was observed in Gr.1 – 1.4% and in Gr.2 – 1,9% (P =0.8402, OR -1,33). In both groups we observed high percent Cesarean section (Gr.1 – 32.3% and Gr.2 – 29.4% (P=0.7651, OR -1,0909), but we did not find a statistical difference between the groups. In both groups, percent of macrosomia was high, despite good glycemic control maintained through pregnancies: 20.0% and 23.0%, for Gr.1 and Gr.2, respectively (P =0.9236, OR -1,0256), and again no statistical difference was found between the groups. Percent of neonatal hypoglycemia was lower in Gr.2 (1.9%), than in Gr.1 (4.41%) (P=0.9236, OR –1,0256).. Percent of respiratory distress syndrome was 2.94% and in 3.92% for Gr.1 and Gr.2, respectively (P= 0.9694, OR – 0.9893), with no statistical difference between the groups.

**Conclusions:** We did not find differences between patients who treated with diet + insulin therapy and with diet + metformin. Percent of preeclampsia, preterm delivery, macrosomia and perinatal death was similar in both groups, only maternal weight gain was lower in the metformin group.

## Introduction

GDM is a widespread complication of pregnancy, defined as glucose intolerance of various severity that is first diagnosed during pregnancy [1]. According to the International Diabetes Federation (IDF) in 2019 “every sixth newborn was born to a mother with hyperglycemia” [2]. In a systematic review published in 2012, the prevalence of GDM varied according to diagnostic criteria, so: American Diabetes Association (ADA) reporting the prevalence of GDM of 2 to 19 percent; Carpenter and Coustan to 3.6 - 38 percent; National Diabetes Data Group: 1.4 - 50 percent; and according to the World Health Organization - 2 - 24.5 percent [4,5]. Women with GDM, detected during pregnancy, are at greater risk of adverse pregnancy outcomes. These include high blood pressure and a baby large for gestational age, that can make a normal birth difficult. Detection of hyperglycemia during pregnancy and the presence of normoglycemia during pregnancy can reduce these risks. The pharmacotherapy options in patients with GDM, who require pharmacotherapy, are insulin or oral antihyperglycemic agents (metformin or glyburide). Some meta-analysis studies have shown that both oral antihyperglycemic agents and insulin therapy can improve pregnancy outcomes in patients with GDM or type 2 diabetes [3].

## The aim

of the study that was carried out at our Center was to assess the efficacy of different treatment options (insulin and metformin) in women with GDM.

## Methods

The study was carried out at the National Center for Diabetes Research in Tbilisi, Georgia in 2015 – 2022. Screening for GDM was performed in 2,422 pregnant women and GDM was revealed in 116 of them. The “One-step” 75-g oral glucose tolerance test (OGTT), derived from the IADPSG (International Association of Diabetes and Pregnancy Study Groups) [5], ADA [23], FIGO (International Federation of Gynecology and Obstetrics) [7] and WHO [6] criteria was used. OGTT was performed at 24-28 weeks of gestation. The test was performed after 8 hour fasting period; fasting plasma glucose was measured, then patients were advised to drink solution that contained 75g of dry glucose powder dissolved in 250ml of lukewarm water, then blood samples were drowned 1 and 2 hours post load. If a plasma glucose value is above or equal to: 92mg/dL (5.1mmol/L) fasting, 180mg/dL (10.0mmol/L) 1hr, and 153mg/dL (8.5mmol/L) 2hr, GDM is suspected. Three patients were referred to our center by antenatal clinics with hyperglycemia, so no OGGT was performed on them: one patient was on the18-th week of gestation (fasting plasma glucose/FPG - 182mg/dl), the second – on the 31-st week (FPG -138mg/dl), and the 3-rd – on the 30-th week of gestation, her FPG was 149mg/dl. As in all three patients FPG levels were above 92 mg/dl (5.1mmol/L), diagnosis of GDM was put before they were admitted to our Center.

After the diagnosis of GDM was confirmed, all necessary clinical and laboratory tests and investigations were performed. Follow-up throughout the pregnancy: adjustment of daily calorie intake - once in trimester; correction of intensive insulin therapy or metformin dose based on the self-blood glucose monitoring (SBGM) – once in a week; registration of hypoglycemia episodes – every visit; HbA1c – every 4 weeks; SBGM – 7 – 6 times a day; microalbuminuria – every two weeks; creatinine – once in trimester; TSH and thyroid peroxidase antibodies - once in trimester; blood pressure control – every visit (antihypertensive therapy was initiated if blood pressure ≥135/85 mmHg); ultrasound examination, cardio monitoring of a fetus, obstetrical/gynecologic follow-up; consultations of a neurologist, cardiologist and nephrologist – once in trimester; low-dose aspirin 75 mg/day from the end of the first trimester.

The patients were divided into 2 groups (Gr.): Gr.1 - 68 patients, who were treated with diet + insulin, and Gr.2 – 51 patients, who were treated with diet + metformin. All patients had their first pregnancy.

Lifestyle behavior change was an important component of the management of GDM in our patients. Treatment started with medical nutrition therapy, physical activity, and weight management, depending on pre-gestational weight. Calorie intake for each patient was calculated individually and depended on the pre-pregnancy body mass index (BMI). BMI of 20-29kg/m^2^ was registered in 86 of our patients, they were recommended a calorie intake of 30-32 kcal\per kg of body weight/day, BMI of 33 patients was 29.5 -33kg/m^2^, and they were recommended 24-25 kcal\ per kg of body weight/daily. Calories were divided as follows: 175g of carbohydrates, 71g of protein, and 28g of fiber. Concerning fats – it was recommended to increase the amount of monounsaturated and polyunsaturated fats, limit saturated fats and exclude trans fats [20,12]. We recommended effective exercise (aerobic, resistance, or both), with its duration of 20–50 min/day (2–7 days/per week of moderate intensity exercise) [21].

Glucose monitoring was aimed at the targets recommended by the 5-th International Workshop-Conference on Gestational Diabetes Mellitus [22]. Following glycemic targets was implemented: fasting BG – 60 - 90 mg/dl (3.3 – 5.0mmol/l), postprandial 1-hours BG < 140mg/dl (<7.7mmol/l), postprandial 2-hours BG < 120mg/dl (< 6.6mmol/l) and BG before each meal – 75-105 mg/dl (4.1-5.8mmol/l). According to the ADA Recommendations the A1C target of <6% (42 mmol/mol) is optimal during pregnancy, if it can be achieved without significant hypoglycemia [23]. SBGM was recommended for all our patients, blood glucose was measured – fasting and postprandial (1- and 2-hours/2-3 times a day); eight of our women were using continuous glucose monitoring (CGM) systems (24).

At the start, (18 -31 gestation weeks), all patients were diet and exercise for a minimum period of one week, and if their glycemic control was unsatisfactory, medication based treatment was initiated. Based on the FIGO recommendations [18,43] insulin therapy was initiated if: age of gestation at GDM diagnosis <20 or >30 weeks; fasting plasma glucose levels >110 mg/dL (6.1mmol/l); 1-hr postprandial glucose >140 mg/dL (7.7mmol/l); pregnancy weight gain >12 kg.

Insulin type and regimens were individualized [18,19]. Rapid-acting and long-acting insulin analogues were used. The starting dose of long-acting insulin analogue (glargine) was 0.2UI/kg/24h, at bedtime. To achieve adequate glycemic control, the doses were adjusted every 3 days. If preprandial glucose levels were normal and postprandial ones were high, rapid-acting insulin analogue (glulizin -0.1 UI/kg/meal) was added before each meal.

In our study, we initial metformin dose - 1000 mg/day (500 mg 2 times a day) and if adequate control (fasting – 60 - 90 mg/dl (3.3 – 5.0mmol/l), postprandial 1-hr < 140mg/dl (<7.7mmol/l), postprandial 2-hr < 120mg/dl (< 6.6mmol/l), before meal – 75-105 mg/dl (4.1-5.8mmol/l) was not achieved, the dose was raised the next week to 1500 mg/day (500 mg 3 times a day), the maximal dose was 2000mg/day. Patients who did not achieve satisfactory glycemic control with metformin received supplemental insulin analogues, Glargine was initiated in 4 women in the 30-32 weeks of gestation, while in all cases metformin was continued at the maximal dose.

The main aims of GDM management are to achieve normoglycemia during all courses of pregnancy, since numerous studies indicate that normal fetal growth can be achieved if in patients with GDM, glucose levels during pregnancy are close to normal [28]. In our study the women were followed up until they delivered and then postpartum.

Definition of some terms: Preterm delivery - birth before 37 weeks of gestation; Preeclampsia - hypertension, edema, and proteinuria after 20 weeks of gestation; Neonatal hypoglycemia - a plasma glucose level of less than 40mg/dl (2.2mmol/l) in the first 48 hours of life; Macrosomia - neonatal weight ≥4,000g.

Statistical analysis: The mean and standard deviation for continuous variables were used to calculate the standardized mean difference (SMD). The odds ratio (OR) was calculated for dichotomous variables with 95% confidence intervals (CIs).

Primary outcomes include a maternal glycemic control (HbA1c) during pregnancy and maternal weight gain at birth. Secondary outcomes included incidence of preeclampsia, preterm delivery, cesarean section, macrosomia, LGA infants (large-for-gestational-age infant, defined as birth weight ≥ 90th percentile for gestational age and infant’s sex), malformations, respiratory distress syndrome and perinatal mortality.

## Results

There were no statistically significant differences between groups 1 and 2 in terms of maternal age, pre-pregnancy BMI, gestational age, and HbA1c levels at baseline (Table 1).

**Table 1.**

Clinical Data of Women 119 with GDM
|  | Gr.1 – 68 | Gr.2 - 51 | P(Gr.1-2) |
| --- | --- | --- | --- |
| Age (years) | 26.2 ± 5.1 | 25.9 ± 5.3 | 0.967536 |
| Pre-pregnancy BMI (kg/m<sup>2</sup>) | 25.6 ± 4.9 | 25.9 ± 5.1 | 0.921329 |
| Gestation age at the beginning of treatment (weeks) | 25.4 ± 4.36 | 25.9 ± 3.98 | 0.932648 |
| HbA1c (%) | 6.5 ± 0.5 | 6.4 ± 0.5 | 0.3876 |
| Microalbuminuria (%) | 5.8 | 5.9 |  |

At the start of the treatment, HbA1c levels for Gr.1 and 2 were 6.5% (0.5) and 6.4% (0.5), respectively. At the 3^rd^ trimester HbA1c levels statistically decreased in both groups: Gr.1 – 6.01% (0.5) and Gr.2 – 5.9% (0.6), while no statistical difference was observed between the groups (P = 0,3239) (Figure 1).

**Figure 1.**
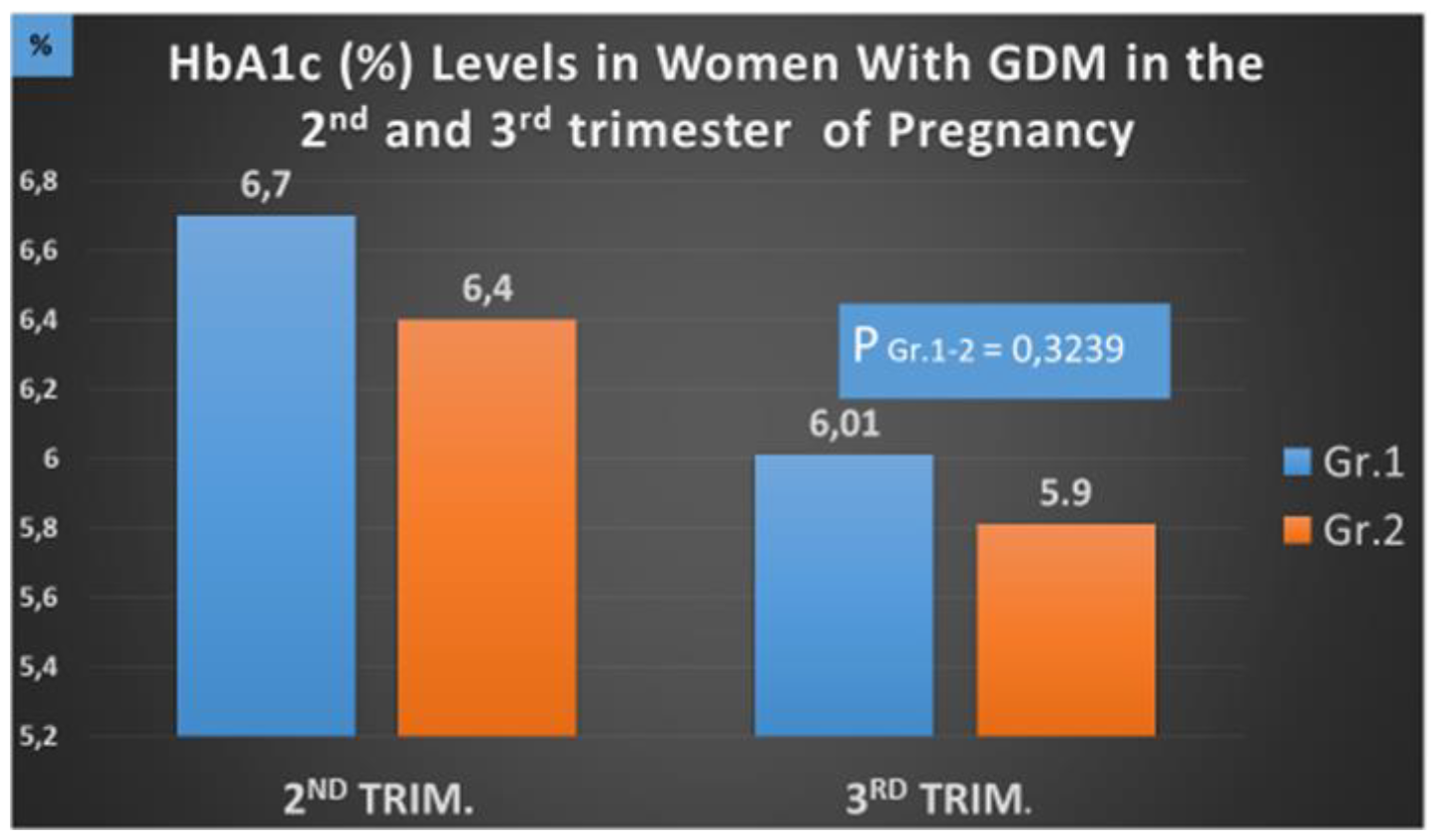
HbA1c levels in women with GDM in the 2nd and 3rd trimester of pregnancy.

From the start of the treatment to before delivery (36–39 gestational weeks), women from Gr.2 (metformin treated group) gained less weight compared to women from Gr.1 (insulin treated group) (1.89±3.88 vs. 4.53±3.67 kg; P=0.003). The same data were obtained in a multicenter, open-label, parallel arms, randomized clinical trial (The Metformin for Gestational Diabetes - 2021), that was performed at 2 hospitals in Málaga (Spain), the studies enrolled women with gestational diabetes who needed pharmacologic treatment [39]. In support of this, recent meta-analyses have shown that weight gain in GDM patients treated with metformin during pregnancy was significantly lower than in those treated with insulin [41,42].

We observed preeclampsia in patients from both groups. In Gr.1 patients, percent of pre-eclampsia was 2.9% and in Gr.2 it was 3.9% (P= 0,7773, OR – 1,33). The insulin-treated group had a lower incidence of preeclampsia compared to the metformin-treated group, but the difference was not significant. Preeclampsia in women with GDM was also observed in other studies. Some studies identify different causes of preeclampsia, as the HAPO study showed that the occurrence of preeclampsia is positively associated with blood glucose levels [8]. In the study conducted by Catalano PM at all., the development of preeclampsia in women with GDM was directly associated with overweight and obesity [9], the same results were obtained in a Swedish retrospective cohort study [14], but, another retrospective cohort study showed that obesity is not associated with the occurrence of preeclampsia in women with GDM [15]. In some retrospective cohort studies, have shown that gestational diabetes, regardless of other causes, leads to the development of preeclampsia [10,11,13,15]. Results obtained in our study have been confirmed by the randomized controlled trial conducted in New Zealand and Australia, the incidence of preeclampsia in the metformin group was lower than in the insulin group, the difference was not statistically significant [16]. Other studies have confirmed the fact, that the appointment of metformin during pregnancy had no effect on the incidence of preeclampsia [17,18,19]. However, in the metformin group, weight gain after the start of treatment was less [16,18].

In our study, percent of preterm deliveries was lower in Gr.1 -4.41%, when compared to Gr.2-5.88% (P=0.7311, OR –1,33), though there was no statistical difference between the groups. The incidence of preterm delivery before 37 weeks of gestation in the insulin-treated group (4.41%) was lower than that in the metformin-treated group (5.88%), but the difference was not statistically significant (P=0.7311, OR –1,33) (Table 2, Figure 2). In our study, the use of metformin did not induce the increase in the incidence of preterm delivery before 37 weeks of gestation. However, results obtained in other studies have shown that gestational diabetes is associated with an increased risk of preterm delivery, birth trauma and respiratory distress syndrome [25,26].

**Table 2.**

Clinical Data of Women with GDM
|  | Gr.1 - 68 | Gr.1 - 51 | P(Gr1-2) | OR |
| --- | --- | --- | --- | --- |
| Preeclampsia | 2 (2.9%) | 2 (3.9%) | 0.7773 | 1.33 |
| No of deliveries Vaginal | 67.6 % | 70.6% |  |  |
| Cesarean section | 32.3% | 29.4% | 0.7651 | 1.0909 |
| Gestational weeks of delivery | 36 - 40 | 36 - 40 |  |  |
| Preterm delivery < 37 weeks | 3 (4.41%) | 3 (5.88%) | 0.7311; | 1.33 |
| Birth weight (g) | $3495 \pm 493.5$ | $3570 \pm 541.3$ | | |

**Fig. 2.**
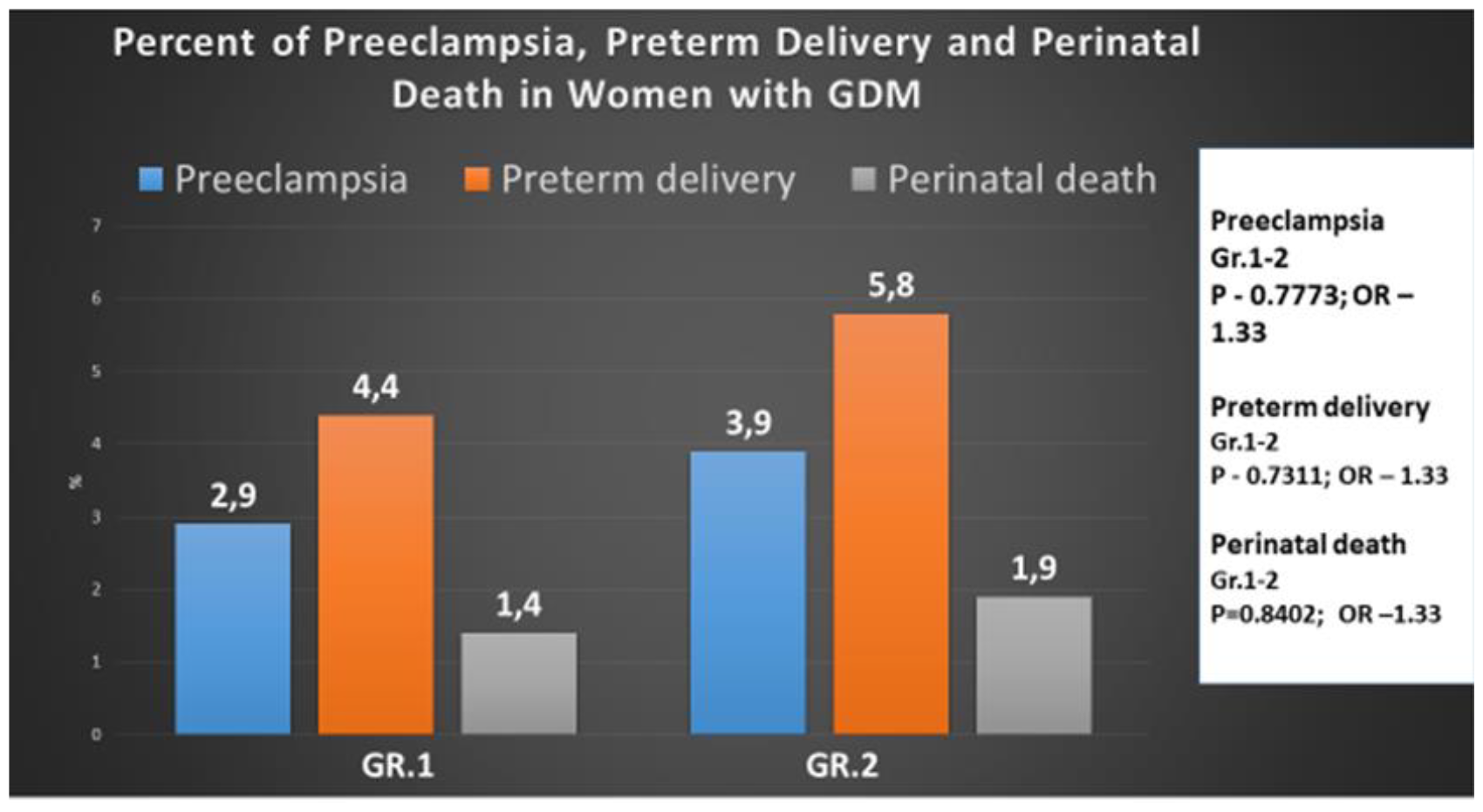
Percent of preeclampsia, preterm delivery and perinatal death in women with GDM.

In our study perinatal mortality was observed in both groups, it comprised 1.4% in Gr.1 and 1.9% in Gr.2 (P =0.8402, OR -1,33), and again, no statistical difference was found between the groups (Figure 2).

Stillbirth was registered only in Gr.1 and its rate was 1.4%, while neonatal death – only in Gr.2 (1.9%) (Table 3). A retrospective study in the United States (more than 4 million women) showed that the risk of stillbirth between 36 and 42 weeks of gestation was higher in women with GDM than in women without GDM (17.1 versus 12.7 per 10,000 births); (RR 1.34; 95% CI 1.2-1.5) [27].

**Table 3.**
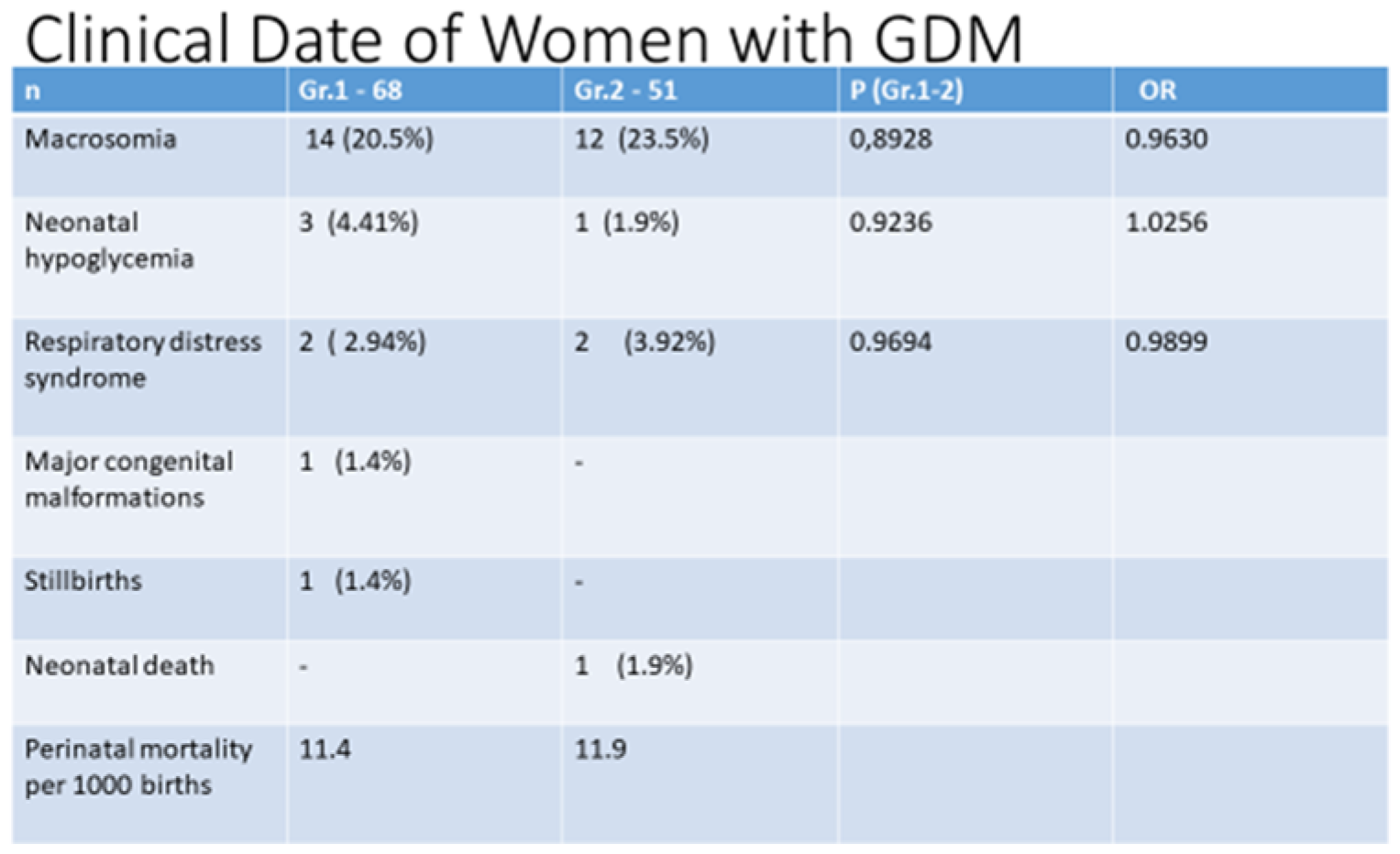

Clinical Date of Women with GDM
| n | Gr.1 - 68 | Gr.2 - 51 | P (Gr.1-2) | OR |
| --- | --- | --- | --- | --- |
| Macrosomia | 14 (20.5%) | 12 (23.5%) | 0,8928 | 0.9630 |
| Neonatal hypoglycemia | 3 (4.41%) | 1 (1.9%) | 0.9236 | 1.0256 |
| Respiratory distress syndrome | 2 (2.94%) | 2 (3.92%) | 0.9694 | 0.9899 |
| Major congenital malformations | 1 (1.4%) | - |  |  |
| Stillbirths | 1 (1.4%) | - |  |  |
| Neonatal death | - | 1 (1.9%) |  |  |
| Perinatal mortality per 1000 births | 11.4 | 11.9 |  |  |

Percent of Cesarean section was high in both groups: Gr.1 – 32.3% and in Gr.2 – 29.4% (P=0.7651, OR -1,0909). However, we did not find a statistical difference between the groups (Table 2). Newborns birth weight was 3495±493. 5g in Gr.1 and 3570±541.3g in Gr.2 (P=0.9186) (Table 2), we did not find a statistical difference between groups.

In both groups, percent of macrosomia was high, despite good glycemic control maintained through pregnancies: 20.0% and 23.0%, for Gr.1 and Gr.2, respectively (P =0.9236, OR -1,0256), and again no statistical difference was found between the groups. Thus, we have found no relationship between the different treatment methods (insulin and metformin) use in patient with GDM and macrosomia development risk (Table 3).

Percent of neonatal hypoglycemia was lower in Gr.2 (1.9%), than in Gr.1 (4.41%) (P=0.9236, OR –1,0256), however we did not find a statistical difference between the groups, either (Table 3). Percent of respiratory distress syndrome was 2.94% and in 3.92% for Gr.1 and Gr.2, respectively (P= 0.9694, OR – 0.9893), with no statistical difference between the groups. Major congenital malformations were observed only in 1 newborn born to a mother from Gr.1.

## Discussion

Until recently, insulin therapy has been the most common treatment for patients with gestational diabetes [30], approximately 50% of women with GDM are prescribed insulin therapy for treating hyperglycemia [29,38], but for the last few years, metformin has been increasingly used in the treatment of GDM [33]. In 2019, was published meta-analysis, that included 28 studies (3976 women with GDM), this study compared the weight of the newborn and different treatments for gestational diabetes: insulin and metformin. It was shown that newborns treated with metformin had a lower birth weight (mean difference - 107.7 g, 95% CI). -182.3 to -32.7), reduced risk of LGA (OR 0.78; 95% CI 0.62-0.99) and macrosomia (OR 0.59; 95% CI 0.46-0.7) than in newborns whose mothers received insulin [34]. In the Australian-New Zealand clinical trial, 751 women with GDM were enrolled to receive either metformin or insulin; researchers did not find significant differences in the results obtained, such as neonatal hypoglycemia, respiratory distress syndrome, hyperbilirubinemia, low Apgar scores, birth trauma and preterm birth [29].

In 2017, a review was published (n = 1487), which included 8 randomized controlled trials evaluating the effects of oral antidiabetic pharmacological therapy (metformin, glyburide and acarbose) in the treatment of pregnant women with GDM, the results showed that the benefits and potential harms of these treatments compared with each other are unclear [35]. Other meta-analyses have compared glibenclamide or metformin with insulin in women with gestational diabetes requiring medical treatment. The following conclusions were made: glibenclamide is clearly inferior to both insulin and metformin, while metformin (plus insulin if needed) works slightly better than insulin. Less frequent episodes of gestational hypertension have been observed in patients taking metformin compared with glibencamide or insulin [36,37], but the use of metformin has been associated with an increased risk of preterm birth compared with insulin [37].

Metformin reduces gluconeogenesis and reduces insulin resistance at the receptor level, it practically cannot cause episodes of hypoglycemia, and also does not lead to weight gain. The results of the use of metformin during pregnancy in patients with polycystic ovary syndrome are well known. Therefore, it can be considered that metformin may be an alternative to insulin in the treatment of gestational diabetes. Another multicenter, open-label, randomized, parallel-group clinical trial conducted in 2 hospitals in Malaga, Spain, evaluated the efficacy of metformin versus insulin in 200 women with GDM and concluded that metformin treatment was associated with better postprandial glycemic control than insulin, lower risk of hypoglycemic episodes, less maternal weight gain, most perinatal outcomes were similar across groups [32].

The primary objective of our study was to evaluate glycemic control in women with GDM treated with metformin or insulin. After the introduction of the insulin and metformin, HbA1c levels statistically decreased in both groups: Gr.1 – 6.01 (0.5) and Gr.2 – 5.9 (0.6) and before delivery we did not find a statistical difference between groups (P = 0,3239). The results of our studies show that insulin, like metformin, effectively controlled glycemia during pregnancy in patients with GDM. In addition, from the start of the treatment before delivery, women from Gr.2 (metformin treated group) gained less weight compared to women from Gr.1(insulin treated group) (1.89±3.88 vs 4.53±3.67 kg; P=0.003). So, maternal weight gain was significantly lower in the metformin group. However, other studies have shown different results from ours, as a randomized prospective study published in 2021 found no trend towards lower weight gain during pregnancy in women in the metformin compared to insulin group [44].

The secondary objective of this study was to assess the incidence of preeclampsia, preterm delivery, cesarean section, macrosomia, malformations, respiratory distress syndrome and perinatal mortality. The incidence of preeclampsia in the insulin-treated group was lower than that in the metformin-treated one, the difference was not statistically significant (P= 0,7773, OR – 1,33). The percentage of preterm deliveries was lower in Gr.1 (insulin-treated group) compared to Gr.2 (metformin-treated group), though there was no statistical difference between the groups. (P=0.7311, OR –1,33). In our study perinatal mortality was observed in both groups, it comprised 1.4% in insulin treated patients and 1.9% in metformin groups, and no statistical difference was found between the groups (P =0.8402, OR -1,33). Stillbirth was registered only in Gr.1 and its rate was 1.4%, while neonatal death – only in Gr.2 (1.9%). Percent of Cesarean section was high in both groups: in insulin treated patients, it was - 32.3% and in metformin treated patients – 29.4%, and no statistical difference between the groups (P=0.7651, OR -1,0909). In both groups, percent of macrosomia was high, despite good glycemic control maintained through pregnancies: 20.0% and 23.0%, for insulin and metformin treated patients respectively, and no statistical difference was found between the groups.

We did not find significant differences between the groups (insulin and metformin) in perinatal outcomes. Our study shows that, the use of metformin in the treatment of patients with gestational diabetes effectively and safely. 119 women to receive either metformin or insulin therapy, finding no significant difference incidence of preeclampsia, preterm delivery, cesarean section, macrosomia, malformations, respiratory distress syndrome and perinatal mortality.

While comparing insulin and metformin therapy, we observed that metformin and insulin are equally effective in the management of gestational diabetes and that metformin was linked to less maternal weight gain (p = 0.003).

## Summary

There was no significant difference in efficacy of metformin and insulin in controlling diabetes in pregnant patients from the two groups, therefore, the efficacy of metformin and insulin was found to be comparable in the management of pregnancy with diabetes in this study. The percent of preeclampsia, preterm delivery, macrosomia, neonatal hypoglycemia, respiratory distress syndrome and perinatal death was similar in both groups. We observed that only maternal weight gain was lower in the metformin group.

## Data Availability

All data produced in the present work are contained in the manuscript

https://doi.org/10.2337/dc23-S015

